## Supplementary Materials for "Estimating the trend of COVID-19 in Norway by combining multiple surveillance indicators"

### Supplementary methods

#### Estimating prevalence from for the "Fraction Positive - Survey" indicator

For the "Fraction Positive - Survey" indicator we want to estimate the symptomatic prevalence of those surveyed, but not everyone with symptoms chose to get tested. We therefore assume that the test positivity rate is equal among all participants who report having cold symptoms and use a Bayesian model to estimate the overall prevalence as follows:

$$p_{\text{covid}} = p_{sympt} * p_{pos}$$
$$N_{sympt} \sim binomial(N, p_{sympt})$$
$$N_{pos} \sim binomial(N_{test}, p_{pos}),$$

where $N$ is the total number of participants, $N_{sympt}$ the number with symptoms, $N_{pos}$ the number of symptomatic participants who took a test and $N_{test}$ the number who tested positive. Using this setup we can estimate the overall symptomatic prevalence $p_{covid}$ with uncertainty.

#### Estimating incidence from Prevalence

Incidence, $I(t)$ and prevalence, $P(t)$ are related by:

$$P(t) = \sum_s p(s)I(t-s)$$

where $p(s)$ is the probability of testing positive for someone who was infected at $s = 0$. We use a PCR-test profile as estimated from [1], which is an approximation for the symptometer data which includes both PCR and antigen tests. To estimate the incidence, we then use an approximate Gaussian process model with a Matern Kernel.

$$logit(I(t)) = i_0 + i(t), i(t) \sim GP(t)$$

.

Similarly following [2], we estimate incidence from wastewater data. Here, the observed amount of SARS-CoV-2 in the wastewater is a convolution of the incidence and the time-profile of gastrointestinal viral load. We assume a standard deviation of 20% of the mean value for the normalised SARS-CoV-2 concentration. We used a similar Gaussian process model as above. For both approaches we use the implementation presented in [3].

#### Disaggregation of weekly to daily data

To disaggregate weekly data, we implement a similar model to the models above where daily infections again are given by

$$logit(I(t)) = i_0 + i(t), i(t) \sim GP(t)$$

.

We then calculate weekly aggregated data $W_i = \sum_t Ind(week = i)I(i)$ where $Ind(week = i)$ is an indicator function for the week. These weekly aggregated incidences are then compared to the aggregated data, $A$

$$A \sim poisson(W).$$

Using the Stan language we can then get samples for $I(t)$ which we then feed into the regression model to ensure we incorporate the uncertainty from the disaggregation.

#### Combining estimated growth rates

We combine the estimated growth rates from the different surveillance systems using methods from the field of meta-analysis where we consider the mean, $r_i(t)$ and the standard deviations $\sigma_i(t)$ calculated from the estimated growth rates from each indicator. We implement a random effects meta-analysis model with a random walk for smoothing over time.

$$r_i(t) \sim normal(\mu_i(t), \sigma_i)$$
$$\mu_i(t) \sim normal(r(t), 0.1)$$
$$r(t = 1) \sim normal(0, 0.1)$$
$$r(t) \sim normal(\alpha + \beta r(t-1), \tau)$$
$$\tau \sim normal(0, 0.5)$$
$$\alpha \sim normal(0, 5)$$
$$\beta \sim normal(0, 5)$$

The usefulness of the combined estimate from all the indicators will depend on the heterogeneity of the individual indicators. We estimate the $I^2$ indicator for the heterogeneity [4] using the following estimate:

$$I^2(t) = \frac{\tau^2(t)}{\tau^2(t) + s^2(t)}$$

where $s^2(t)$ is the geometric mean of the variances from the growth rates of the individual indicators. All the models are implemented in the Stan probabilistic programming language.

#### Estimating the Effective Reproduction Number from the Growth Rate

From the growth rates one can estimate reproduction numbers [5] if we assume a gamma distributed generation with shape, $\alpha$ and rate $\beta$.

$$R = \left(1 + \frac{r}{\beta}\right)^{\alpha}.$$

Following the results in [6] we update the parameters in the gamma distribution by the dominating variant in Norway. For each variant (Wuhan, Alpha, Delta and Omicron) we use a mean given by [6]. To be able to estimate the Reproduction Number we then also need the spread of the distribution, here characterised by the coefficient of varition, $cv = \sigma/\mu$ . For the ancestral Wuhan variant, we use a coefficient of variation of 1.2 [7], and then 0.73 for Alpha, 0.7 for Delta [8] and 0.75 for Omicron [9]. We model the transition between different variants with a logistic function matched to the data on Variants from ECDC [10]. The transition is then given by:

$$f(t) = \frac{1}{1 + \exp\left(\frac{t-d}{w}\right)},$$

where $f$ is the fraction of infections with the new variant, $d$ is the date when the variant reached 50% and $w$ is the width of the transition. $d$ and $w$ were manually extracted from the data. The value of the mean or the coefficient of variation $X_i$ is then given by:

$$X_i(t) = fX_i^n + (1 - f)X_p,$$

where $X_i^n$ is the value for the new variant and $X_i^p$ is the value for the old variant.

#### Relative incidence

From an estimated growth rate, $r(t)$, we can also estimate an associated relative incidence by

$$I(t) = \prod_{i=1}^{t} exp(r(i)).$$

We sample from the estimates of $r(t)$ to quantify the uncertainty in $I(t)$

### Supplementary Results

To investigate the effect of changing the amount of smoothing when estimating growth rates, we have estimated growth rates with different window lengths, $2l + 1$, in the negative binomial regression model, In Figure 1 we show the estimated growth rate for the number of reported cases with changing length of the time-window. This clearly illustrates how a longer time window has lower uncertainty and fewer temporal features. Increased smoothing also shrinks the growth rates towards 0.

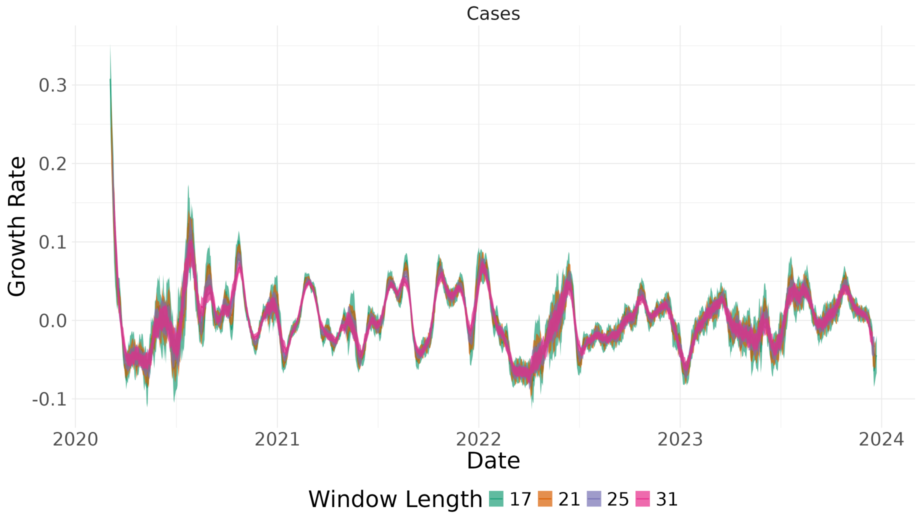


Figure 1: Estimated growth rates for the number of positive cases with varying amount of smoothing controlled by the length of the time-window used, Norway 2020-2023.

Following the same procedures as in the main paper we show in Figure 2, with a window length of 21 days, and Figure 3, with a window length of 31 days, the correlation matrix between the growth rates of the different indicators when we vary the amount of smoothing. Higher smoothing leads to a higher correlation, likely due to two main effects. Since some of our indicators are smoothed by default, for example due to being weekly indicators, it is possible that more smoothing of daily indicators allows us to have more similar amounts of smoothing. Secondly, since smoothing has a tendency to shrink estimates towards 0, one can likely also get some spurious correlations by smoothing too much.

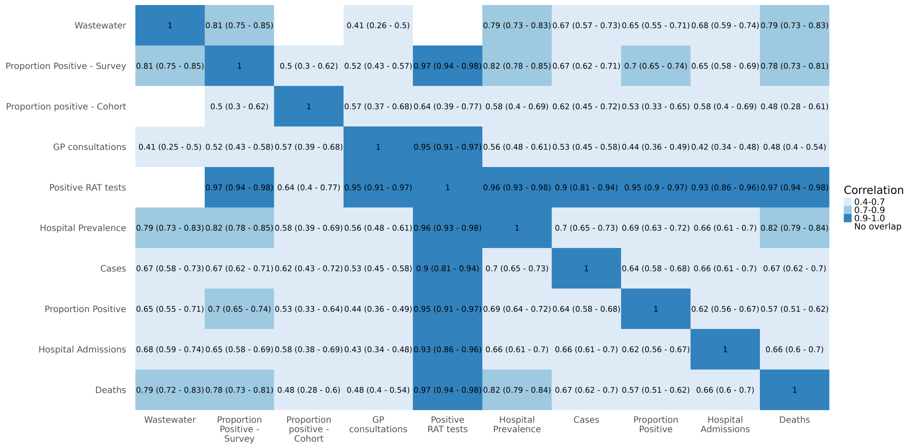


Figure 2: Estimated correlation with 95% confidence intervals between the estimated growth rates of the different surveillance indicators for a smoothing time-window of 21 days, Norway 2020-2023.

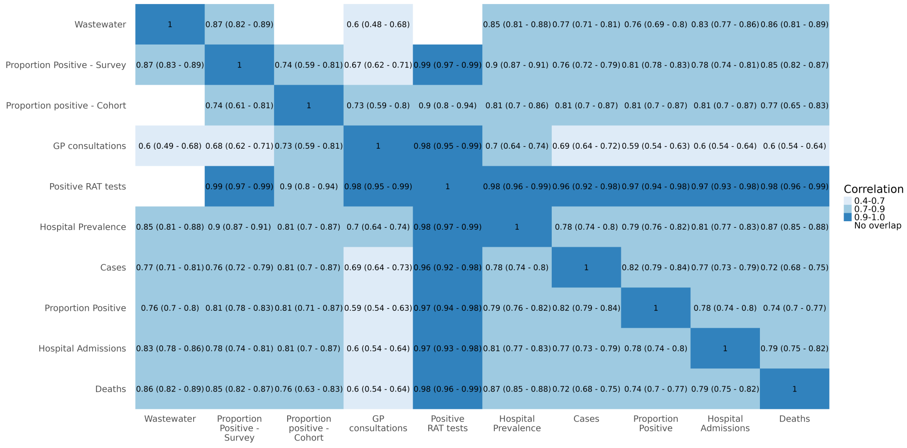


Figure 3: Estimated correlation with 95% confidence intervals between the estimated growth rates of the different surveillance indicators for a soothing time-window of 31 days, Norway 2020-2023.

10. European Centre for Disease Prevention and Control. European Respiratory Virus Surveillance Summary (ERVISS), 2024, Week 17.
